# Polygenic Risk Score predicts QTc-prolongation and Short-Term Mortality in Patients using QT-prolonging Psychoactive Medications

**DOI:** 10.1101/2024.07.24.24310940

**Authors:** Mays Altaraihi

## Abstract

**Background:** There is a genetic component to the QT-interval. This study investigated whether a polygenic risk score for QTc (PRS_QTc_) could predict ΔQTc and short-term mortality in first-time users of QT-prolonging medications (QTPM) with a known risk of Torsade de Pointes.

**Methods:** First-time users of psychoactive QTPM in the Copenhagen Hospital Biobank and the Danish Blood Donor Study from 2009-2021 were included. ΔQTc was calculated and all-cause 30-day mortality following initiation of treatment was explored. All models were adjusted for conventional QT-prolonging risk factors, and models investigating death were additionally adjusted for potential comorbidity confounders.

**Results:** The PRS_QTc_ could predict ΔQTc (2.88 milliseconds (ms) for every increase of standard deviation in PRS_QTc_ (P <0.001)) following treatment initiation. Individuals in the top ≥ 80 % of PRS_QTc_ had a higher risk of ΔQTc of ≥60 ms compared to individuals in <80 % PRS_QTc_ (OR = 4.88 P = 0.019). Furthermore, the study has also shown that the shorter QTc before initiation of QTPM, the higher the risk of greater ΔQTc.

A high PRS_QTc_ could also predict short-term mortality following treatment initiation: Individuals in the top PRS_QTc_ ≥90 % had an odds ratio of 1.84 (P-value = 0.002) for short-term mortality compared to individuals with PRS_QTc_ <90 %. Individuals in the top PRS_QTc_ ≥99 % had an odds ratio of 4.95 (P-value = 0.009) for short-term mortality compared to individuals in the <99 % PRS_QTc_

It could be replicated that PRS_QTc_ ≥90 % was a predictor of short-term mortality with OR 1.52 (P-value = 0.002) compared to PRS_QTc_ <90 % in a bigger cohort (N=15.249).

**Conclusion:** PRS_QTc_ seems to be predictive of ΔQTc following initiation of treatment. PRS_QTc_ proves to be a sufficient predictor of 30-day mortality after initiation of QT-prolonging psychoactive drugs with a known risk of Torsade de Pointes.

If used in a clinical setting, PRS_QT_ may help prevent sudden cardiac deaths associated with QTPM.

## Introduction

The electrocardiographic QT-interval represents the sum of the ventricular depolarization and repolarization. The QT-interval is clinically important as both a long and a short QT-interval predisposes to ventricular arrhythmias and sudden cardiac death. Many commonly prescribed drugs (QT-prolonging medication, QTPM) may cause QT-interval prolongation and thereby increase the risk of developing Torsade de Pointes (TdP) which predisposes to sudden cardiac death (1–3). Therefore, evaluation of drug-induced QT-prolongation is crucial when developing, testing, and prescribing drugs.

The mechanisms of drug-induced long QT involve the drug’s interaction with several cardiac ion channels, in which the main interaction is a blockade of the rapid potassium current (I_Kr_) encoded by the gene *KCNH2*. The final manifestation of the drug-ion-channel interactions is a reduction of the net repolarization current in the ventricular action potential, leading to QT-prolongation (4,5). Another important risk factor of QT-prolongation is hypokalemia(6,7). Not all patients develop prolonged QT-interval when exposed to the same QTPM and this variability in response is partly ascribed to genetic factors (8). Indeed, studies show that variants in genes encoding the cardiac channels can modify the drug-channel interaction, resulting in an increase of the binding affinity or the channel gaiting kinetics (9,10). Genome-wide association studies (GWAS) of the QT-interval have been conducted to explain the heritability of the QT-interval in the general population. Studies show that polygenic risk scores calculated based on GWASs of corrected QT-interval (PRS_QTc_) associate significantly with long QT-syndrome in genotype negative individuals and sudden cardiac death (2,11).

This study hypothesized that PRS_QTc_ may predict the variability in response to QTPM. By designing a case-control study, the aim of the current study was to investigate if individuals in the top quantile of PRS_QTc_ are at greater risk of prolonged QT-interval following initiation of QT-prolonging psychoactive medication with a known risk of TdP. Subsequently, the aim was also to investigate whether individuals belonging in the top quantile of PRS_QTc_ are at greater risk of 30-day-mortality following the initiation.

## Methods

### Study Design and Population

The study is based on the Copenhagen Hospital Biobank (CHB)(12) initiated in February 2009 and the Danish Blood Donor Study (DBDS)(13) initiated in March 2010. First-time users of psychoactive drugs listed on CredibleMeds (14) as QTPM with a known risk of TdP from 2009-2021 were included. These drugs will hereby be referred to as reference drugs. The following reference drugs were investigated: Citalopram, Haloperidol, Escitalopram, Methadone, Chlorprothixene, Levomepromazine, Pimozide, Sulpiride and Sertindole. For inclusion, the drugs had to be tablets or capsules intended for oral administration.

Twelve lead ECGs were obtained per clinical indication and integrated in the CHB/DBDS environment. Based on the Marquette 12SL algorithm used in the ECGs, ECGs not consistent with a valid measurement of the QT-interval (e.g., branch blocks, atrial fibrillation, bradycardia, see Supplementary for ECG details) were excluded.

Apart from being first-time users of reference drugs, included individuals had to be registered with ECGs with valid measurements of the QT-interval. When exploring the outcome, ΔQTc, individuals had to be registered with at least two valid ECGs: At least one ECG measured within 18 months before the first prescription of the reference drug (baseline-ECG) and at least one ECG measured within 18 months after the first prescription (treatment-ECG). If more than one baseline-ECGs were present, the ECG closest to the date of the first prescription was used. If more than one after-ECGs were present, the ECG closest to day 15 after the first prescription was used. Furthermore, only individuals with after-ECGs, in which the ECG was measured within the treatment period, were included. For more details on calculation of the treatment period, see Supplementary. When investigating short-term mortality, only a valid baseline-ECG was required to be included.

Lastly, using the latest blood test, all included individuals had to be registered with a potassium blood test within 4 months before the first prescription of the reference drug. Figure 1 and Figure 2 illustrate the methods used.

**Figure 1:**
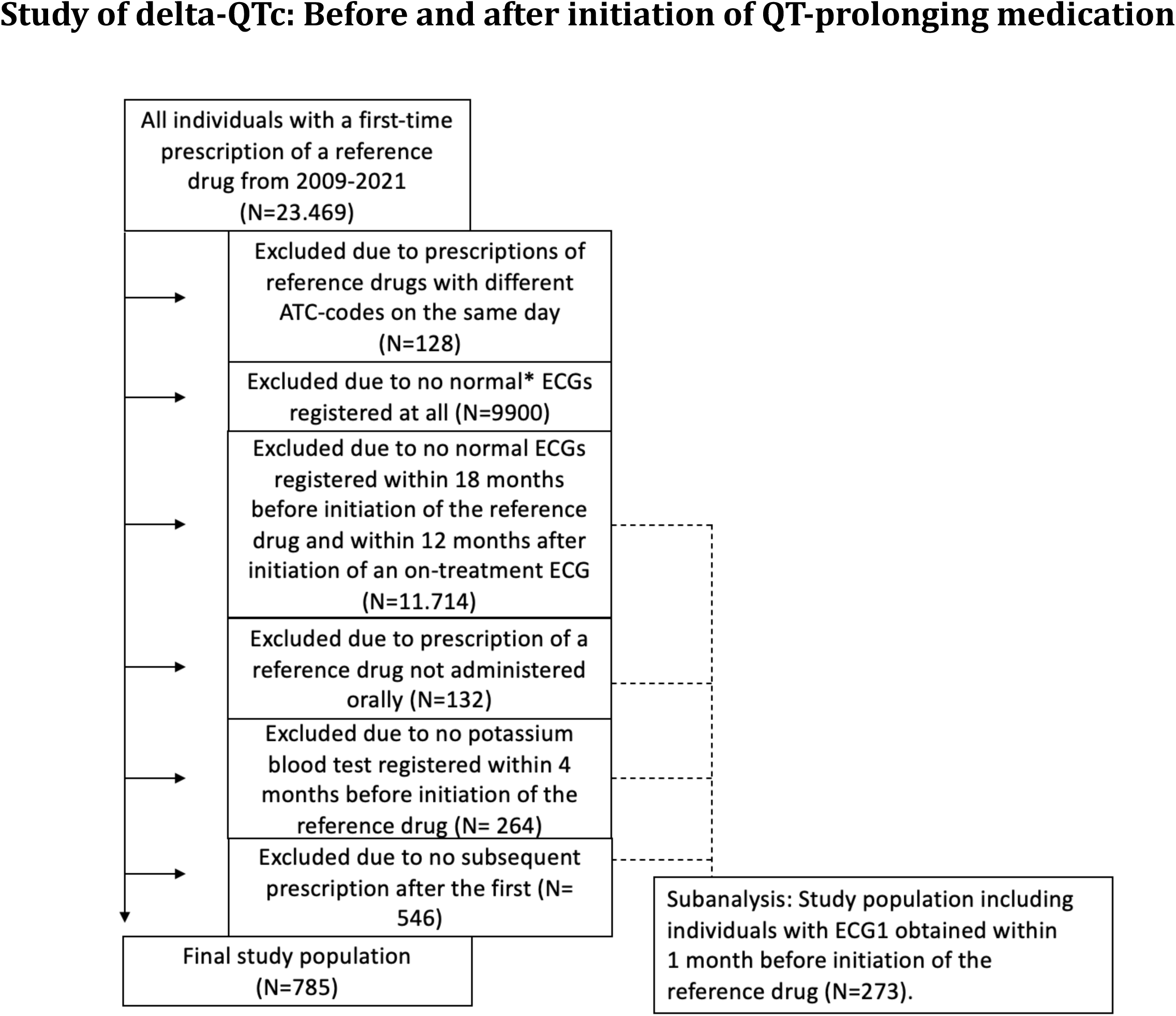
Selection of the study population with a baseline ECG and on-treatment ECG along with a baseline potassium blood test. *Normal ECG: No presence of atrial Aibrillation/Alutter, right or left bundle branch block, pace rhythms, 2. or 3. degree AV block, premature atrial or ventricular extrasystoles, Wolf-Parkinson-White pattern, or poor-quality data. We additionslly excluded ECGs with a QRS duration >120 ms, bradycardia (heart frequency, <40) or tachycardia (heart frequency, >110). We also excluded extreme outliers based on the following criteria: QTcFridericia (QTcF) <0.1 % of cohort distribution (<320 milliseconds), and QTcF >99.9 % (>614 milliseconds).

**Figure 2:**
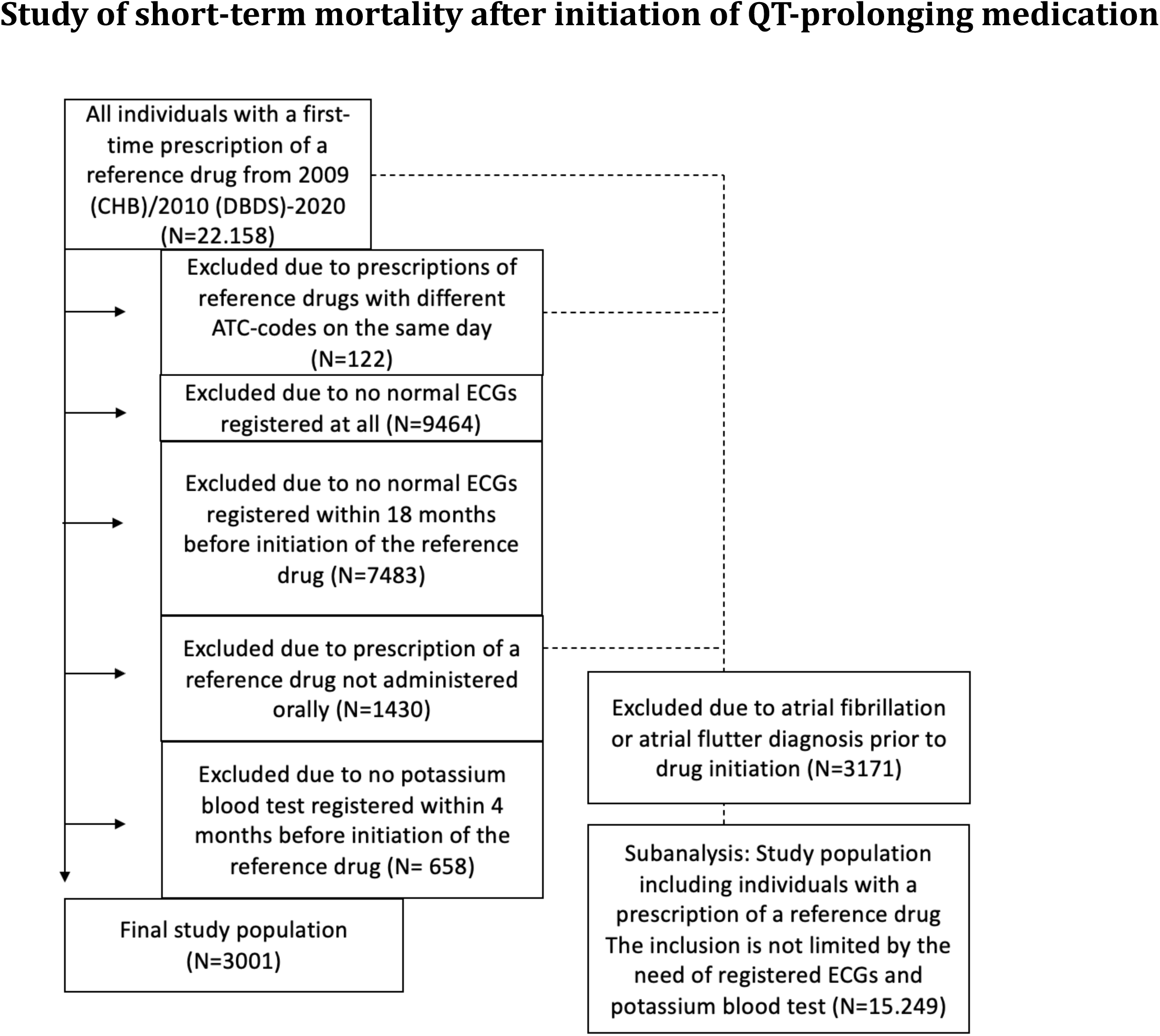
Selection of the study population investigating the outcome: On-treatment, 30-day-mortality.

### Genetics

GWAS summary statistics on Fridericia-corrected QT-interval (QTcF) from previous published data were queried (15). Using PRS-CS (16), individual polygenic risk in CHB/DBDS was calculated. In CHB/DBDS, individuals were genotyped on Infinium Global Screening Array (Illumina Inc, USA). In sample-wise quality control of CHB/DBDS, samples with missingness < 97%, sex mismatch, genetic ancestry outliers and heterozygosity outliers (>3SD) were removed. In marker-wise QC, variants with missingness < 99%, low Hardy-Weinberg equilibrium (P < 0.0001), and minor allele frequency < 0.1% were removed. Imputed genetic data with an information quality cutoff < 0.9 were used to create genotype files for individuals in CHB/DBDS.

### Other covariates

For model adjustment, the following conventional risk factors for prolonged QT-interval were investigated:

Age. Sex. Hypokalemia (plasma-Potassium <3.6 mmol/L). Loop-diuretics or thiazides without potassium: If prescribed within 30 days before prescription of the reference drug had been filled, the individuals were registered as being on diuretic treatment (17).

To explore the additive effect of QTPM on the outcome, it was investigated whether the individuals had a prescription of another QT-prolonging psychoactive medication with a *possible* risk of Torsade de Pointes according to CredibleMeds (14). The prescription of the other QTPM had to be within 30 days before the prescription of the reference drug. The additional QT-prolonging psychoactive medication could be one of the following drugs: Mirtazapine, Nortriptyline, Misanserine, Fluanxol, Clozapine, Aripiprazole, Perphenazine, Melperone, Maprotiline and Pipamperone.

Models were adjusted for heart failure, using all diagnosis codes for heart failure. When addressing ischemic heart disease (IHD), both disease diagnoses and/or procedure codes involved in ischemic heart diseases were used: Ischemic myocardial infarction diagnoses (IHD) percutaneous coronary intervention or coronary artery bypass grafting (18).

Inhibited drug clearance due to hepatic or renal dysfunction can prolong the QT-interval when exposed to QTPM. Therefore, hepatic or renal insufficiency before prescription of the reference drug was included as a covariate. Diagnosis codes for hepatic failure chronic hepatitis, fibrosis/cirrhosis of the liver, acute kidney failure, chronic kidney disease or unspecified kidney failure were used (19).

For short-term mortality, the models were adjusted for the most common mortality causes in high-income countries: Cancer, IHD, stroke and chronic obstructive pulmonary disease (COPD). Cancer was defined as all cancers except non-malignant melanoma skin cancer. All the used diagnosis and procedure codes had to be registered before the prescription of the reference drug. All the ATC, diagnosis and procedure codes can be found in Supplementary.

### Outcomes

The outcome, ΔQTc, was defined as the absolute difference of measured QTcF in treatment-ECG and baseline-ECG. The outcome, ΔQTc, was investigated in a linear regression model adjusted for covariates. The outcome, ΔQTc, was also investigated as a binary outcome in adjusted logistic models, using ΔQTc ≥10 ms, ≥20 ms, ≥30 ms, ≥40 ms, ≥50 ms and ≥60 ms. The binary ΔQTc outcomes were investigated in an interaction analysis, exploring the interaction between PRS_QTc_ and the QTcF in the baseline-ECG.

Short-term mortality was defined as death within 30 days after the first filled prescription of the reference drug. The death had to occur during the treatment period. In addition, only deaths registered to be of natural causes were registered as outcomes, excluding suicides, homicides, accidents, and unknown causes. The decision to use 30-day-mortality as outcome is based on the risk of drug-induced TdP being greatest during the following days after initiation of QTPM (18).

To address ascertainment bias, short-term mortality was tested as outcome in a cohort not limited by the need of ECGs and potassium blood tests prior to initiation of the reference drug. In this cohort, all individuals with the diagnosis atrial fibrillation or atrial flutter were excluded, as atrial fibrillation reduces the likelihood of TdP after initiation of QTPM(20).

### Statistics

The case-control association analysis was performed using linear and logistic regression with the generalized linear model (GLM). The results are shown as ΔQTc in milliseconds or odds ratios with belonging confidence intervals and P-values. When calculating statistically significant differences of continuous outcomes in separate groups, the Mann-Whitney U test was used. If the outcomes were binary, Chi-squared test was used. In the latter two tests, only the P-values are shown in this study.

## Results

### ΔQTc

The cohort exploring the outcome, ΔQTc, consisted of 785 first-time users of QTPM meeting the inclusion criteria. This study found that for every increase in standard deviation (s.d.) of PRS_QTc_, ΔQTc would increase with 2.88 ms (P <0.001) (table 1 and figure 3).

**Table 1:**
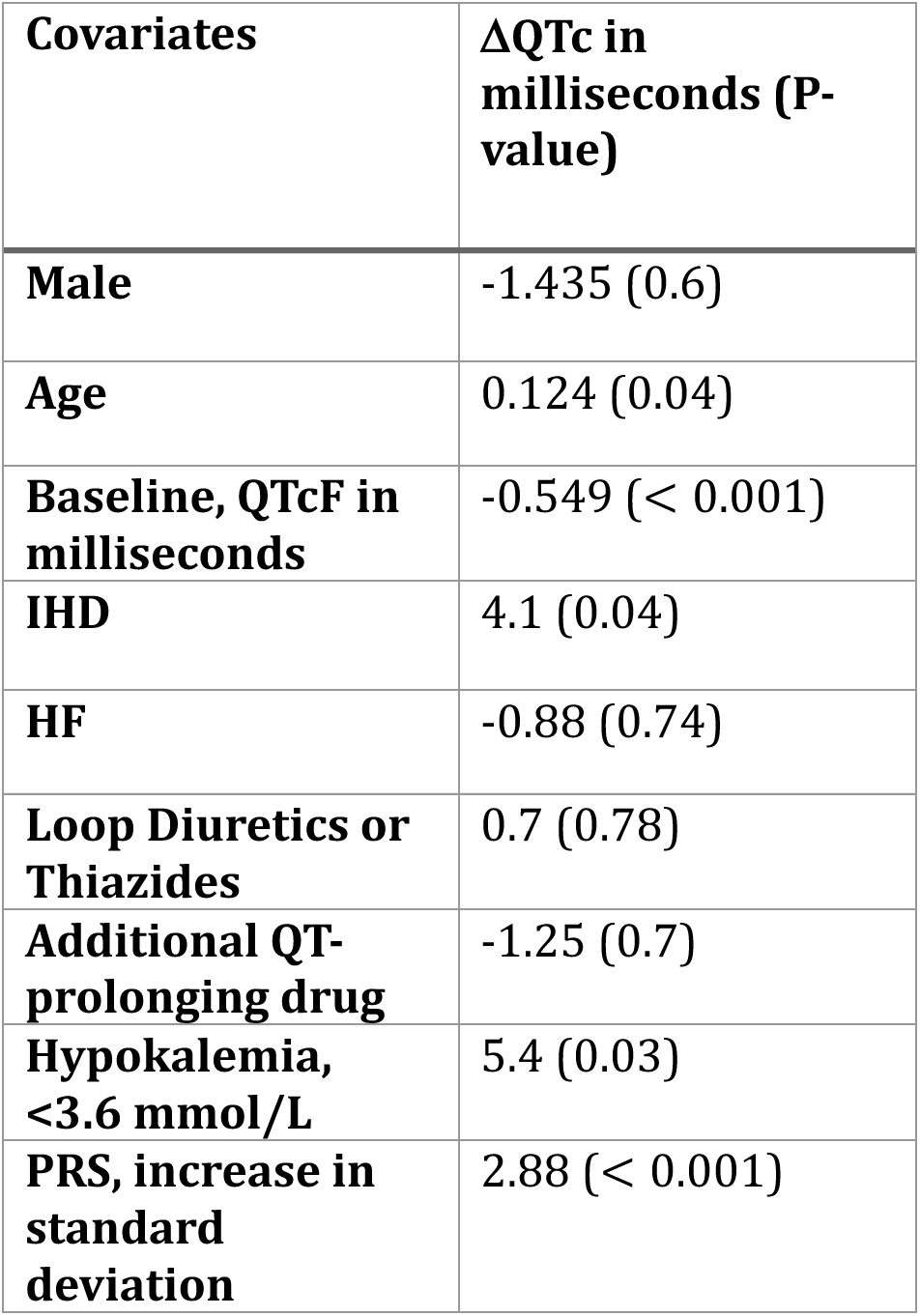
The absolute delta-QTcF in milliseconds of ECGs measured within 18 months before initiation of QT-prolonging medication and within 12 months after initiation of on-treatment ECG. The inAluence of different QT-prolonging factors on delta-QTcF are shown. The linear model used have additionally been adjusted for PCs 1-10.

**Figure 3:**
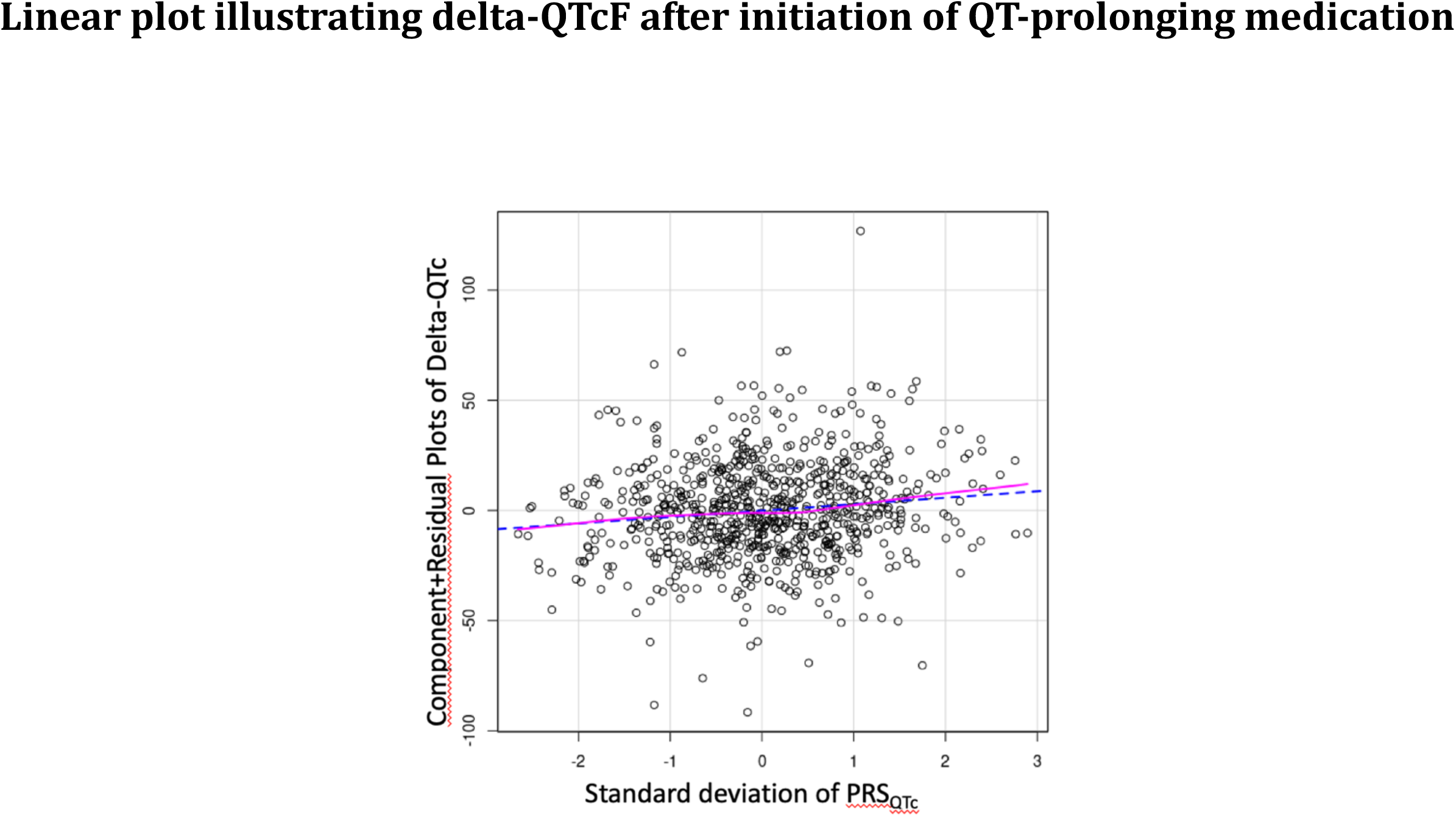
Adjusted linear plot showing the absolute delta-QTcF of ECGs measured before and after initiation of QT-prolonging drugs as function of PRS_QTc_. The plot illustrates the study population in which ECG1 is measured within 18 months before initiation (N=785). ECG2 are on-treatment ECGs obtained within 12 months after initiation of the QT-prolonging drug. The continuous line represents the component plot, and the dashed line represents the residual plot.

The binary traits ΔQTc ≥10 ms, ≥20 ms, ≥30 ms, ≥40 ms, ≥50 ms and ≥60 ms (table 2) were examined. Individuals in the top ≥80 % PRS_QTc_ had greater risk of ΔQTc ≥60 ms compared to individuals with PRS_QTc_ <80 % (OR = 4.88, P = 0.019). Results from the interaction analysis showed that individuals with normal QTc in the baseline-ECG (380-449 ms (M)/390-459 ms (F)) were at greatest risk of higher ΔQTc compared to those with borderline QTc (QTcF 450-496 ms (M)/460-479 ms) or prolonged QTc (470 ms (M)/ ≥ 480 ms (F)) in the baseline-ECG (table).

**Table 2:** Results showing the OR of the outcomes, delta-QTc: >=10 ms, >=20 ms, >=30 ms, >=40 ms, >=50 ms and >=60 ms, from logistic models including variables potentially inAluencing delta-QT. All the logistic models used have additionally been adjusted for PCs 1-10.

| **Factors influencing delta QT interval (N=785).** | **≥ 10 ms,**<br>**OR (P-value)** | **≥ 20 ms,**<br>**OR (P-value)** | **≥ 30 ms,**<br>**OR (P-value)** | **≥ 40 ms,**<br>**OR (P-value)** | **≥ 50 ms,**<br>**OR (P-value)** | **≥ 60 ms,**<br>**OR (P-value)** |
| --- | --- | --- | --- | --- | --- | --- |
| **Male** | 0.89 (0.48) | 0.81 (0.24) | 0.93 (0.74) | 1.33 (0.36) | 1.41 (0.4) | 1.39 (0.55) |
| **Age** | 1.01 (0.01) | 1 (0.24) | 1.01 (0.31) | 1.02 (0.14) | 1.03 (0.12) | 1.03 (0.28) |
| **Baseline, QTcF in milliseconds** | 0.97 (< 1 * 10<sup>-4</sup>) | 0.96 (< 1 * 10<sup>-4</sup>) | 0.96 (< 1 * 10<sup>-4</sup>) | 0.96 (< 1 * 10<sup>-4</sup>) | 0.95 (< 1 * 10<sup>-4</sup>) | 0.93 (< 1 * 10<sup>-4</sup>) |
| **IHD** | 1.41 (0.07) | 1.4 (0.11) | 1.37 (0.23) | 1.76 (0.089) | 2.09 (0.09) | 6.02 (0.003) |
| **HF** | 1.18 (0.52) | 1.12 (0.69) | 1.11 (0.78) | 1.26 (0.6) | 1.33 (0.61) | 1.31 (0.72) |
| **Loop Diuretics or Thiazides** | 1.54 (0.08) | 1.19 (0.513) | 1.05 (0.88) | 1.56 (0.27) | 1.25 (0.69) | 1.07 (0.93) |
| **Additional QT-prolonging drug** | 1.11 (0.73) | 1.01 (0.97) | 0.62 (0.3) | 0.15 (0.07) | 0.35 (0.34) | 0.31 (0.37) |
| **Hypokalemia, <3.6 mmol/L** | 1.1 (0.68) | 1.14 (0.16) | 1.62 (0.12) | 2.72 (0.007) | 3.82 (0.003) | 9.38 (0.00014) |
| **PRS ≥ 80th percentile** | 1.95 (0.00075) | 2.11 (0.0005) | 1.84 (0.02) | 2.36 (0.015) | 2.4 (0.06) | 4.88 (0.019) |
| **PRS ≥ 90th percentile** | 1.48 (0.15) | 1.91 (0.03) | 1.39 (0.4) | 1.39 (0.56) | 1.38 (0.68) | 1.93 (0.56) |
| **PRS ≥ 99th percentile** | 2.05 (0.35) | 4.54 (0.047) | 0 (0.97) | 0 (0.99) | 0 (0.99) | 0 (0.99) |
| **PRS ≤ 20th percentile** | 0.75 (0.17) | 0.72 (0.16) | 0.52 (0.04) | 0.79 (0.54) | 0.82 (0.69) | 0.86 (0.83) |

Table 2: Results showing the OR of the outcomes, delta- $QTc$ : $\geq 10$ ms, $\geq 20$ ms, $\geq 30$ ms, $\geq 40$ ms, $\geq 50$ ms and $\geq 60$ ms, from logistic models including variables potentially influencing delta- $QT$ .
| $\Delta QTc$ | $\geq 10$ ms,<br>OR (P-value) | $\geq 20$ ms,<br>OR (P-value) | $\geq 30$ ms,<br>OR (P-value) | $\geq 40$ ms,<br>OR (P-value) | $\geq 50$ ms,<br>OR (P-value) | $\geq 60$ ms,<br>OR (P-value) |
| --- | --- | --- | --- | --- | --- | --- |
| **PRS ≥ 80th percentile** | 1.9 (0.001) | 2.07 (<0.001) | 1.78 (0.03) | 2.6 (0.004) | 2.3 (0.076) | 5.9 (0.015) |
| **PRS ≥ 90th percentile** | 1.56 (0.00075) | 2.04 (0.017) | 1.4 (0.4) | 1.77 (0.27) | 1.44 (0.64) | 2.28 (0.48) |
| **PRS ≥ 99th percentile** | 2.19 (0.3) | 4.7 (0.041) | 1.84 (0.02) | 0 (0.98) | 0 (0.98) | 0 (0.99) |

The study has also shown that the shorter QTc before initiation of QTPM, the higher the risk of greater ΔQTc (figure 4). This is in line with an Icelandic study showing that the shorter QTc before surgery, the higher the risk of greater ΔQTc postoperative (21).

**Figure 4:**
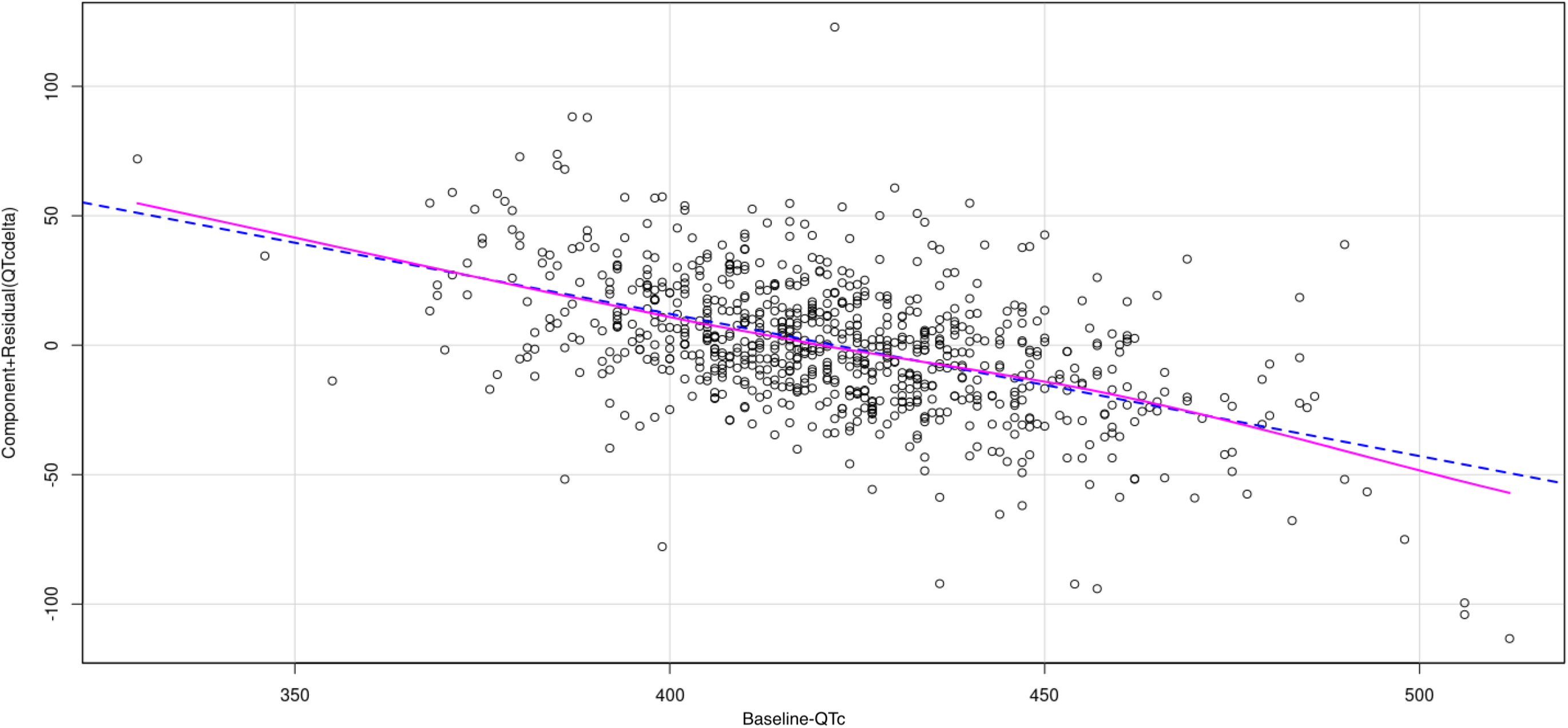
This linear plot illustrates that the shorter QTc before initiation of QT-prolonging drugs, the higher the risk of greater ΔQTc after initiation of treatment.

### Short-term mortality

In the studied population consisting of 3001, PRS_QTc_ ≥80 % could predict short-term mortality with OR 1.32 (P-value = 0.089), PRS_QTc_ ≥90 % could predict short-term mortality with OR 1.84 (P-value = 0.002) and PRS_QTc_ ≥99 % could predict short-term mortality with OR 4.95 (P-value = 0.0093) (table 5). All the reported results originate from adjusted models including PCs 1-10, age, sex, heart failure, IHD, loop diuretics/thiazides, additional QTc-prolonging medication, renal insufficiency, hepatic insufficiency, hypokalemia, baseline QTcF, cancer, stroke and COPD.

**Table 3:** Analyzing the interaction of PRS_QTc_ >=80 % and intervals of QTcF from ECGs measured within 18 months before initiation, using the binary outcome: Delta-QTc: >=10 ms, >=20 ms, >=30 ms, >=40 ms, >=50 ms and >=60 ms. The ECGs used after initiation have all been on-treatment and within 12 months after initiation. The models have been adjusted for PCs 1-10.

| **Baseline QTcF in ms** | **Interaction P-value, $\geq \Delta 10$ ms** | **Interaction P-value, $\geq \Delta 20$ ms** | **Interaction P-value, $\geq \Delta 30$ ms** | **Interaction P-value, $\geq \Delta 40$ ms** | **Interaction P-value, $\geq \Delta 50$ ms** | **Interaction P-value, $\geq \Delta 60$ ms** |
| --- | --- | --- | --- | --- | --- | --- |
| **Normal QTc: 380-449 (M), 390-459 (F)** | 0.01 | 0.059 | 0.034 | 0.079 | 0.16 | 0.0098 |
| **Borderline QTc: 450-469 (M), 460-479 (F)** | 0.28 | 0.78 | 0.46 | 0.99 | 1 | 1 |
| **Long QTc: $\geq 470$ (M), $\geq 480$ (F)** | 0.14 | 0.15 | 0.31 | 0.31 | 1 | 1 |

**Table 4:**
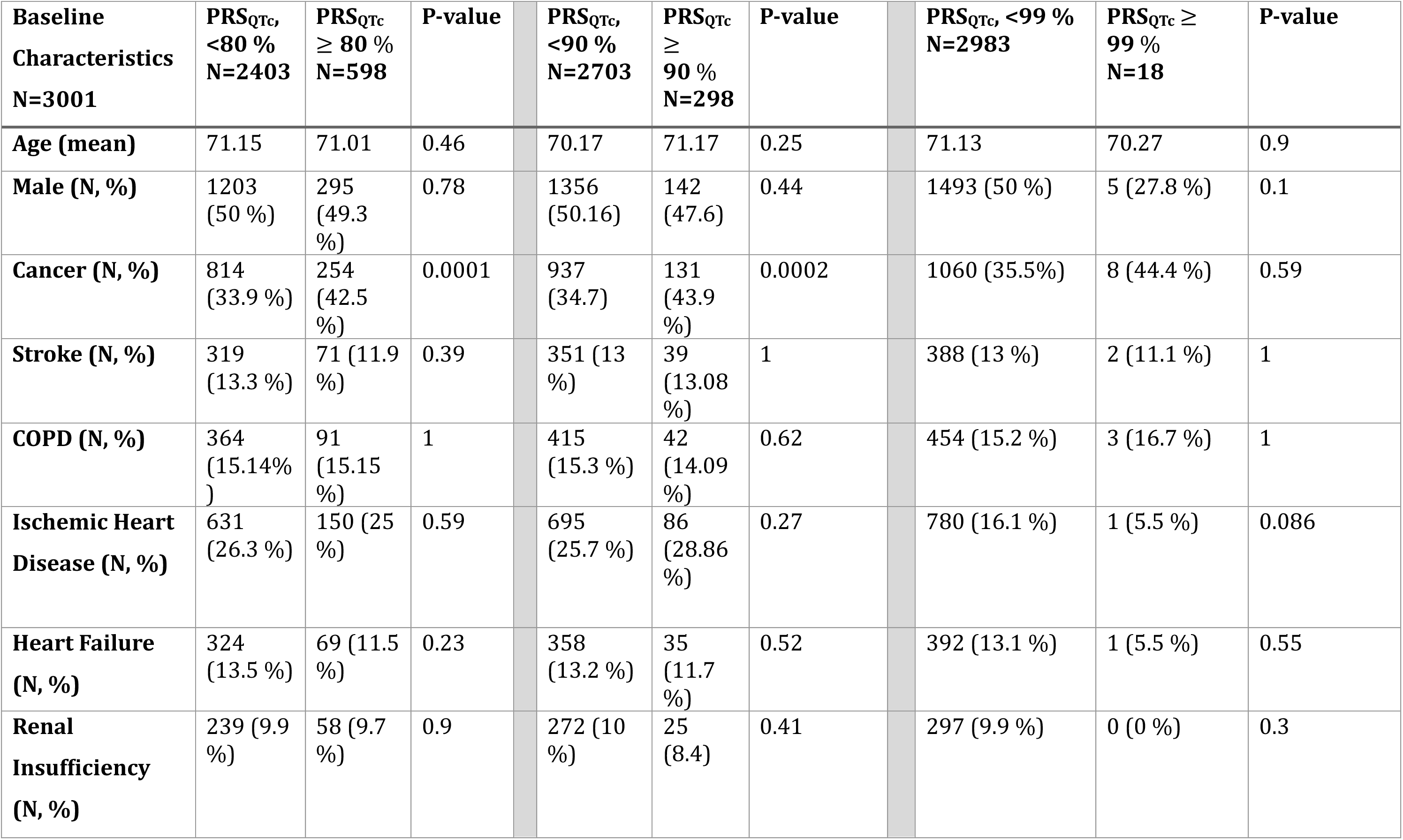
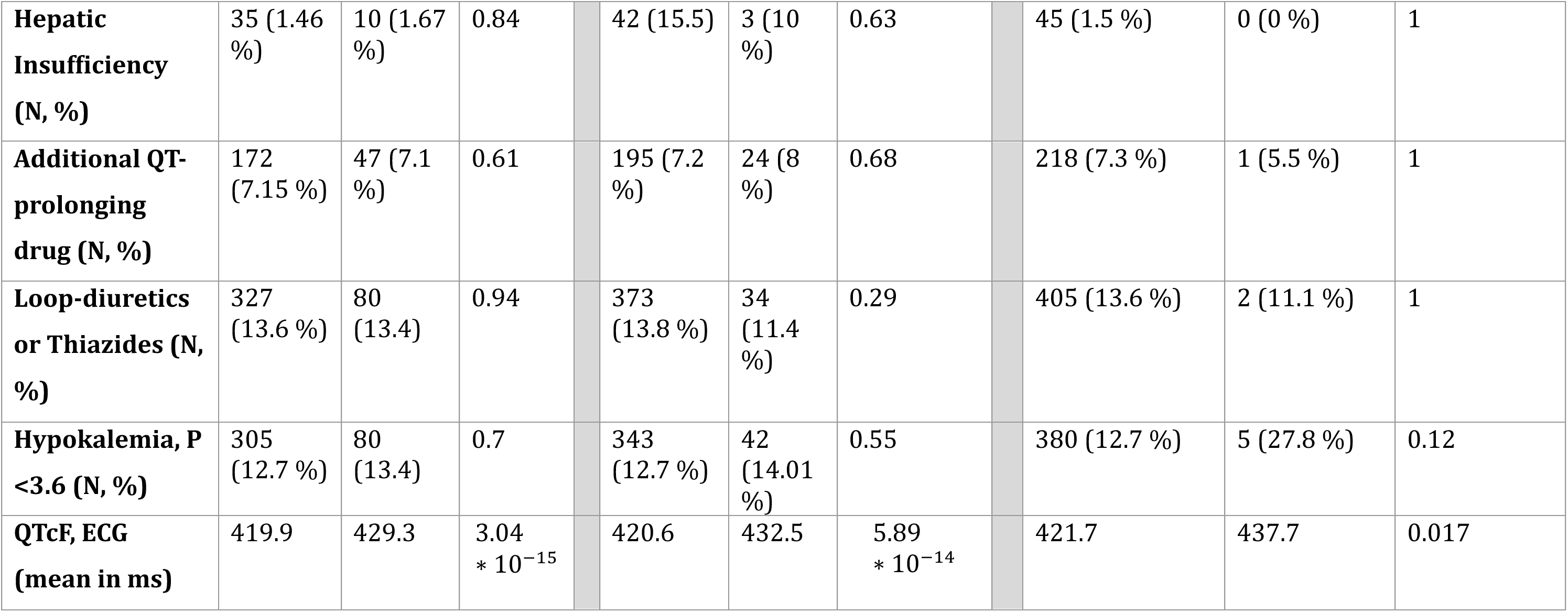
The baseline characteristics of the study population used to calculate OR of 30-day-mortality after initiation of the QT-prolonging medication.

| **Baseline Characteristics**<br>**N=3001** | **PRS<sub>QTc</sub>,**<br>**<80 %**<br>**N=2403** | **PRS<sub>QTc</sub>**<br>**≥ 80 %**<br>**N=598** | **P-value** |  | **PRS<sub>QTc</sub>,**<br>**<90 %**<br>**N=2703** | **PRS<sub>QTc</sub>**<br>**≥ 90 %**<br>**N=298** | **P-value** |  | **PRS<sub>QTc</sub>, <99 %**<br>**N=2983** | **PRS<sub>QTc</sub> ≥ 99 %**<br>**N=18** | **P-value** |
| --- | --- | --- | --- | --- | --- | --- | --- | --- | --- | --- | --- |
| **Age (mean)** | 71.15 | 71.01 | 0.46 |  | 70.17 | 71.17 | 0.25 |  | 71.13 | 70.27 | 0.9 |
| **Male (N, %)** | 1203<br>(50 %) | 295<br>(49.3 %) | 0.78 |  | 1356<br>(50.16) | 142<br>(47.6) | 0.44 |  | 1493 (50 %) | 5 (27.8 %) | 0.1 |
| **Cancer (N, %)** | 814<br>(33.9 %) | 254<br>(42.5 %) | 0.0001 |  | 937<br>(34.7) | 131<br>(43.9 %) | 0.0002 |  | 1060 (35.5%) | 8 (44.4 %) | 0.59 |
| **Stroke (N, %)** | 319<br>(13.3 %) | 71 (11.9 %) | 0.39 |  | 351 (13 %) | 39<br>(13.08 %) | 1 |  | 388 (13 %) | 2 (11.1 %) | 1 |
| **COPD (N, %)** | 364<br>(15.14% ) | 91<br>(15.15 %) | 1 |  | 415<br>(15.3 %) | 42<br>(14.09 %) | 0.62 |  | 454 (15.2 %) | 3 (16.7 %) | 1 |
| **Ischemic Heart Disease (N, %)** | 631<br>(26.3 %) | 150 (25 %) | 0.59 |  | 695<br>(25.7 %) | 86<br>(28.86 %) | 0.27 |  | 780 (16.1 %) | 1 (5.5 %) | 0.086 |
| **Heart Failure (N, %)** | 324<br>(13.5 %) | 69 (11.5 %) | 0.23 |  | 358<br>(13.2 %) | 35<br>(11.7 %) | 0.52 |  | 392 (13.1 %) | 1 (5.5 %) | 0.55 |
| **Renal Insufficiency (N, %)** | 239 (9.9 %) | 58 (9.7 %) | 0.9 |  | 272 (10 %) | 25<br>(8.4) | 0.41 |  | 297 (9.9 %) | 0 (0 %) | 0.3 |
| **Hepatic Insufficiency (N, %)** | 35 (1.46 %) | 10 (1.67 %) | 0.84 |  | 42 (15.5) | 3 (10 %) | 0.63 |  | 45 (1.5 %) | 0 (0 %) | 1 |
| **Additional QT-prolonging drug (N, %)** | 172 (7.15 %) | 47 (7.1 %) | 0.61 |  | 195 (7.2 %) | 24 (8 %) | 0.68 |  | 218 (7.3 %) | 1 (5.5 %) | 1 |
| **Loop-diuretics or Thiazides (N, %)** | 327 (13.6 %) | 80 (13.4) | 0.94 |  | 373 (13.8 %) | 34 (11.4 %) | 0.29 |  | 405 (13.6 %) | 2 (11.1 %) | 1 |
| **Hypokalemia, P <3.6 (N, %)** | 305 (12.7 %) | 80 (13.4) | 0.7 |  | 343 (12.7 %) | 42 (14.01 %) | 0.55 |  | 380 (12.7 %) | 5 (27.8 %) | 0.12 |
| **QTcF, ECG (mean in ms)** | 419.9 | 429.3 | $3.04 * 10^{-15}$ | | 420.6 | 432.5 | $5.89 * 10^{-14}$ | | 421.7 | 437.7 | 0.017 |

**Table 5:** Results of on-treatment, 30-day-mortality in the study population of Airst-time users of QT-prolonging medications. In the logistic models, the risk models in the left column were included. In addition, all the models have been adjusted for PCs 1-10.

| **Risk factors for mortality and prolonged QT-interval** | **OR for 30-day-mortality** | **Confidence Interval** | **P-value** |
| --- | --- | --- | --- |
| **Age** | 1.02 | 1.01-1.04 | 0.00018 |
| **Male** | 1.7 | 1.28-2.25 | 0.0002 |
| **Cancer** | 6.44 | 4.77-8.69 | $<1*10^{-4}$ |
| **Stroke** | 1.2 | 0.81-1.77 | 0.36 |
| **COPD** | 1.24 | 0.87-1.76 | 0.23 |
| **Ischemic Heart Disease** | 0.94 | 0.68-1.29 | 0.7 |
| **Heart Failure** | 1.02 | 0.67-1.55 | 0.9 |
| **Renal Insufficiency** | 1.07 | 0.7-1.65 | 0.75 |
| **Hepatic Insufficiency** | 2.95 | 1.32-6.62 | 0.0008 |
| **Additional QT-prolonging drug** | 1.2 | 0.73-1.94 | 0.47 |
| **Loop-diuretics or Thiazides** | 1.97 | 1.4-2.79 | 0.0001 |
| **Hypokalemia (P <3.6)** | 1.74 | 1.24-2.45 | 0.0013 |
| **QTcF, ECG** | 0.99 | 0.99-1 | 0.0029 |
| **PRS<sub>QTc</sub> ≥ 80 %** | 1.32 | 0.96-1.81 | 0.089 |
| **PRS<sub>QTc</sub> ≥ 90 %** | 1.84 | 1.26-2.74 | 0.002 |
| **PRS<sub>QTc</sub> ≥ 99 %** | 4.95 | 1.48-16.56 | 0.0093 |

**Table 6:**
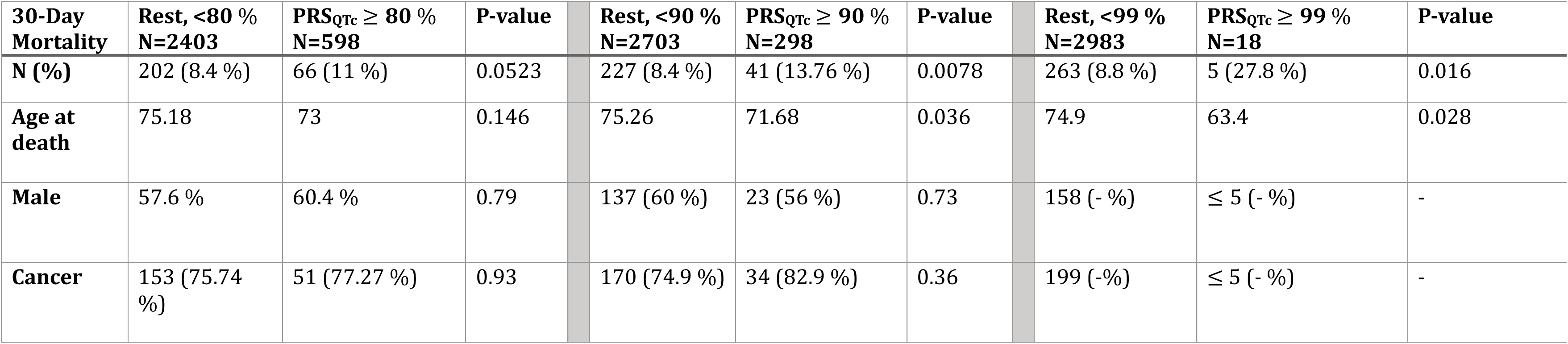
Results showing characteristics of the individuals deceased within 30-days after initiation of the QT-prolonging in the study population.

| **30-Day Mortality** | **Rest, <80 %**<br>**N=2403** | **PRS<sub>QTc</sub> ≥ 80 %**<br>**N=598** | **P-value** |  | **Rest, <90 %**<br>**N=2703** | **PRS<sub>QTc</sub> ≥ 90 %**<br>**N=298** | **P-value** |  | **Rest, <99 %**<br>**N=2983** | **PRS<sub>QTc</sub> ≥ 99 %**<br>**N=18** | **P-value** |
| --- | --- | --- | --- | --- | --- | --- | --- | --- | --- | --- | --- |
| **N (%)** | 202 (8.4 %) | 66 (11 %) | 0.0523 |  | 227 (8.4 %) | 41 (13.76 %) | 0.0078 |  | 263 (8.8 %) | 5 (27.8 %) | 0.016 |
| **Age at death** | 75.18 | 73 | 0.146 |  | 75.26 | 71.68 | 0.036 |  | 74.9 | 63.4 | 0.028 |
| **Male** | 57.6 % | 60.4 % | 0.79 |  | 137 (60 %) | 23 (56 %) | 0.73 |  | 158 (- %) | ≤ 5 (- %) | - |
| **Cancer** | 153 (75.74 %) | 51 (77.27 %) | 0.93 |  | 170 (74.9 %) | 34 (82.9 %) | 0.36 |  | 199 (-%) | ≤ 5 (- %) | - |

A forest plot showing AUC of the different risk factors for prolonged QTc/mortality is illustrated in figure 5. When analyzing the AUC difference of all risk factors except PRS_QTc_ ≥ 90 % and all risk factors including PRS_QTc_ ≥ 90 %, the difference is statistically significant, though not robust, with a P-value of 0.04.

**Figure 5:**
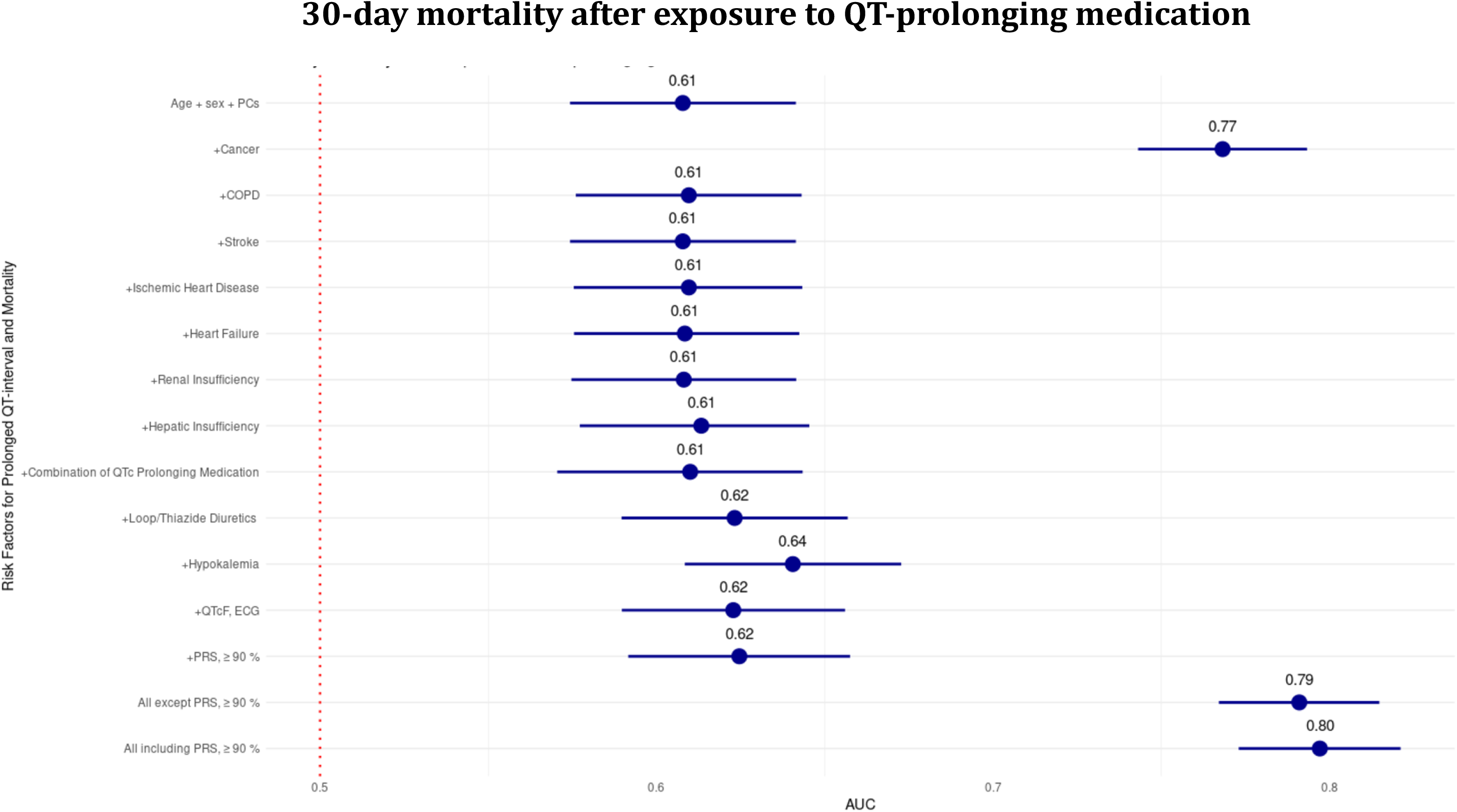
Forest plot showing the AUC of risk factors for mortality and/or prolonged QT-interval after initiation of QT-prolonging medication. The AUC difference of “*All except PRS_QTc_* ≥ 90 %*”* and “*All including PRS_QTc_* ≥ 90 %*”* is statistically signiAicant, though not robust, with a P-value of 0.04.

This study also found that longer baseline QTcF seems to be protective of 30-day-mortality following initiation of the reference drug: QTcF, OR 0.99 (P-value = 0.0029).

When analyzing the deceased individuals, the ones in the top PRS_QTc_ quantiles are slightly younger than the rest: For instance, individuals belonging to the PRS_QTc_ ≥ 99 %, the mean age at death 64.3 years and individuals belonging to PRS_QTc_ < 99 %, the mean age at death was 74.9 years (P-value = 0.028) (table 7).

Another finding is the increased prevalence of cancer in the top PRS_QTc_ quantiles (table 5): For instance, the prevalence of cancer in PRS_QTc_ ≥80 % is 42.5 % and 33.9 % in the PRS_QTc_ < 80 % group (P-value = 0.0001). In the deceased individuals, cancer was not significantly more prevalent in the top PRS_QTc_ quantiles (table 7).

In the sensitivity analysis without the cohort-limiting factors as baseline ECGs and potassium blood tests, the cohort consisted of 15.675 individuals, and the baseline characteristics are shown in Supplementary, table 1. In adjusted models of the sensitivity analysis, PRS_QTc_ ≥80 % could predict short-term mortality with OR 1.23 (P-value = 0.053), PRS_QTc_ ≥90 % could predict short-term mortality with OR 1.52 (P-value = 0.002) and PRS_QTc_ ≥99 % could predict short-term mortality with OR 1.69 (P-value = 0.12) (table 2 in Supplementary).

When analyzing the deceased in this cohort, it was not observed that the individuals in the top PRS_QTc_ quantiles had lower ages at death (table 3 in Supplementary). When investigating the prevalence of cancer in the top PRS_QTc_ quantiles in this cohort, this study observed increased prevalence in the top quantiles: The prevalence of cancer in PRS_QTc_ ≥80 % is 42.5 % and 33.9 % in the PRS_QTc_ < 80 % group (P-value = 0.0001). In the deceased individuals, cancer was not significantly more prevalent in the top PRS_QTc_ quantiles (table 3, Supplementary).

## Discussion

A case-control study investigating whether PRS_QTc_ is of predictive value to the outcome ΔQTc QTc and short-term mortality in first-time users of QT-prolonging psychoactive medication with a known risk of TdP was conducted. This study found that individuals in the top quantiles of PRS_QTc_ may be at risk of greater positive ΔQTc and 30-day-mortality following initiation of treatment.

An abstract presented in the American Heart Association in November 2023 of a prospective cohort study investigating the predictive value of PRS_QTc_ to the outcome delta-QTc in individuals exposed to Sotalol found similar results (22).

It is interesting to discuss the predictive value of PRS_QTc_ in relation to a baseline ECG measured prior to initiation of QTPM. Our results show that the more prolonged the QT-interval is in a baseline ECG, the lower the risk of greater ΔQTc, -0.54 ms (< 1 ∗ 10^-4^). For instance, for every increase in baseline QTcF, the odds of ≥ 60 ms in ΔQTc decrease by a factor 0.93 (P-value = < 1 ∗ 10^-4^). This finding prompts that baseline ECGs cannot predict the individuals at risk of greater QT-prolongation when exposed to QTPM. In contrast, when belonging to the top ≥ 80 % in PRS_QTc_, the odds of ≥ 60 ms in delta-QTcF increase by a factor 4.88 (P-value 0.019). In comparison, when affected by hypokalemia, the odds of ≥ 60 ms in delta-QTcF increase by a factor of 9.38 (P-value = 0.00014).

Results from the interaction analysis show that individuals in the top ≥ 80 % PRS_QTc_ with a normal baseline QTcF were the individuals at greatest risk of positive dΔQTc when exposed to the QT-prolonging psychoactive drugs.

There is no QTc threshold at which TdP is certain to develop, albeit the risk of TdP increases by a twofold to threefold when having a QTc greater than 500 ms. It is also documented that every 10-millisecond increase in QTc, increases the risk of TdP by 5-7 % (23–25).

Based on the results of this study, PRS_QTc_ may be a better predictive tool than the baseline ECG when assessing how an individual will respond to QTPM, especially in individuals with normal baseline QTc.

When stating that PRS_QTc_ may predict 30-day-mortality following initiation of QT-prolonging psychoactive medication, it is important to analyze the baseline characteristics of the studied population. As a baseline ECG and potassium blood test were inclusion criteria in the studied population, most of the individuals originated from the CHB-cohort, which consist of hospitalized patients or outpatients at hospitals. The baseline characteristics of the studied population indicate high frequency of one or more disorders (table 5). This is observed across all PRS_QTc_ groups, though the only variables which are distributed significantly different in the PRS_QTc_ groups are baseline QTcF and cancer: For example, in the group, PRS_QTc_ ≥ 90 %, the mean QTcF was 432.5 ms and the mean QTcF in the <90 % was 420.6ms (P-value= 5.89 ∗ 10^-14^). The prevalence of cancer in top PRS_QTc_ ≥ 90 % was 43.9 % and in the <90 % group, the prevalence was 34.7 % (P-value = 0.0002). It has not been previously reported that individuals with long QT syndrome are at increased risk of cancer. However, GWASs of the QT-interval typically find common variants in genes regulating ion-channels, calcium-handling proteins and myocyte structure(11,15,26,27). Both ion channels and calcium-handling proteins are important drivers for hallmarks of cancer as proliferation, apoptosis and migration (28–31). As genomic instability is an important feature of cancer, it is plausible that some variants influencing the phenotype of the QT-interval may also influence the risk of cancer.

Though there is an increased prevalence of cancer in the top quantiles in the studied population, there is not a significant increased prevalence of cancer in the top PRS_QTc_ quantiles of the deceased individuals. In models adjusted for all risk factors, PRS_QTc_ ≥ 80 % predicts short-term mortality with OR 1.32 (P-value = 0.089), PRS_QTc_ ≥ 90 % predicts short-term mortality with OR 1.84 (P-value = 0.002) and PRS_QTc_ ≥ 99 % predicts short-term mortality with OR 4.95 (P-value = 0.0093).

In the sensitivity analysis including a younger cohort with fewer counts of disorders, PRS_QTc_ ≥ 90 % could still predict short-term mortality with OR 1.52 (P-value = 0.002).

When finding that PRS_QTc_ may both predict the QTc response and short-term mortality following initiation of QTPM, using PRS_QTc_ as a predictive tool in a clinical setting may help prevent sudden deaths associated with QTPM.

One limitation of this study is the time limit at which the baseline ECG and potassium blood test had to be measured within, as they cannot fully represent the biological conditions in all individuals right before the initiation of the QTPM. Another limitation is that, though, the treatment periods based on the prescriptions were calculated, it cannot be fully validated that the individuals were under influence of the reference drugs when the treatment-ECGs were measured. The main strength of this study is the combination of both clinical, paraclinical and genetic variables in cohorts of reasonable sizes, allowing us to compare baseline ECG and PRS_QTc_ to each other, which has led to one of the novel interesting findings.

In conclusion, this study found that PRS_QTc_ can be predictive of the QTc response following initiation of QT-prolonging psychoactive drugs with a known risk of Torsade de Pointes. PRS_QTc_ might even be a better predictor of the QTc response compared to the baseline ECG. This study also found that PRS_QTc_ could predict 30-day-mortality after initiation of treatment. If used in a clinical setting, PRS_QT_ may help prevent sudden cardiac deaths associated with QT-prolonging medication.

## Supporting information

The absolute delta-QTcF in milliseconds of ECGs measured within 18 months (table 1a) and 1 month (table 1b) before initiation of non-QT-prolonging med

## Data Availability

All data produced in the present study are available upon reasonable request to the authors.

## Acknowledgements

I would like to thank Jonas Ghouse, Søren Albertsen-Rand, Henning Bundgaard, Alex Hørby Christensen and Morten Salling Olesen. I would also like to thank deCODE genetics for the work done in Copenhagen Hospital Biobank and the Danish Blood Donor Study.

## References

1. Simpson TF, Salazar JW, Vittinghoff E, Probert J, Iwahashi A, Olgin JE, et al. Association of QT-Prolonging Medications With Risk of Autopsy-Defined Causes of Sudden Death. JAMA Intern Med [Internet]. 2020 May 1 [cited 2023 Nov 1];180(5):698–706. Available from: https://jamanetwork.com/journals/jamainternalmedicine/fullarticle/2762577

2. Al-Akchar M, Siddique MS. Long QT Syndrome. StatPearls [Internet]. 2022 Dec 26 [cited 2024 Jan 7]; Available from: https://www.ncbi.nlm.nih.gov/books/NBK441860/

3. Straus SMJM, Kors JA, De Bruin ML, Van Der Hooft CS, Hofman A, Heeringa J, et al. Prolonged QTc interval and risk of sudden cardiac death in a population of older adults. J Am Coll Cardiol [Internet]. 2006 Jan 17 [cited 2024 Jan 7];47(2):362–7. Available from: https://pubmed.ncbi.nlm.nih.gov/16412861/

4. Klotzbaugh RJ, Martin A, Turner JR. Drug-induced proarrhythmia: Discussion and considerations for clinical practice. JAAPA [Internet]. 2020 Feb 1 [cited 2023 Nov 1];33(2):1–7. Available from: https://pubmed.ncbi.nlm.nih.gov/31990841/

5. El-Sherif N, Turitto G, Boutjdir M. Acquired long QT syndrome and torsade de pointes. Pacing Clin Electrophysiol [Internet]. 2018 Apr 1 [cited 2023 Nov 1];41(4):414–21. Available from: https://pubmed.ncbi.nlm.nih.gov/29405316/

6. Weiss JN, Qu Z, Shivkumar K. Electrophysiology of Hypokalemia and Hyperkalemia. Circ Arrhythm Electrophysiol [Internet]. 2017 Mar 1 [cited 2023 Nov 2];10(3):675. Available from: https://www.ahajournals.org/doi/abs/10.1161/CIRCEP.116.004667

7. Kannankeril P, Roden DM, Darbar D. Drug-Induced Long QT Syndrome. Pharmacol Rev [Internet]. 2010 Dec [cited 2023 Nov 20];62(4):760. Available from: /pmc/articles/PMC2993258/

8. Mahida S, Hogarth AJ, Cowan C, Tayebjee MH, Graham LN, Pepper CB. Genetics of congenital and drug-induced long QT syndromes: current evidence and future research perspectives. Journal of Interventional Cardiac Electrophysiology 2013 37:1 [Internet]. 2013 Mar 21 [cited 2023 Nov 1];37(1):9–19. Available from: https://link.springer.com/article/10.1007/s10840-013-9779-5

9. Abbott GW, Sesti F, Splawski I, Buck ME, Lehmann MH, Timothy KW, et al. MiRP1 Forms IKr Potassium Channels with HERG and Is Associated with Cardiac Arrhythmia. Cell. 1999 Apr 16;97(2):175–87.

10. Sesti F, Abbott GW, Wei J, Murray KT, Saksena S, Schwartz PJ, et al. A common polymorphism associated with antibiotic-induced cardiac arrhythmia. Proceedings of the National Academy of Sciences [Internet]. 2000 Sep 12 [cited 2023 Nov 1];97(19):10613–8. Available from: https://www.pnas.org/doi/abs/10.1073/pnas.180223197

11. Young WJ, Lahrouchi N, Isaacs A, Duong TV, Foco L, Ahmed F, et al. Genetic analyses of the electrocardiographic QT interval and its components identify additional loci and pathways. Nature Communications 2022 13:1 [Internet]. 2022 Sep 1 [cited 2024 Jan 7];13(1):1–18. Available from: https://www.nature.com/articles/s41467-022-32821-z

12. Sørensen E, Christiansen L, Wilkowski B, Larsen MH, Burgdorf KS, Thørner LW, et al. Data Resource Profile: The Copenhagen Hospital Biobank (CHB). Int J Epidemiol [Internet]. 2021 Jun 1 [cited 2023 Nov 2];50(3):719–720E. Available from: https://pubmed.ncbi.nlm.nih.gov/33169150/

13. Erikstrup C, Sørensen E, Nielsen KR, Bruun MT, Petersen MS, Rostgaard K, et al. Cohort Profile: The Danish Blood Donor Study. Int J Epidemiol [Internet]. 2023 Jun 6 [cited 2024 Jul 23];52(3):e162–71. Available from: 10.1093/ije/dyac194

14. Home :: Crediblemeds [Internet]. [cited 2024 Jan 7]. Available from: https://www.crediblemeds.org/

15. Hoffmann TJ, Lu M, Oni-Orisan A, Lee C, Risch N, Iribarren C. A large genome-wide association study of QT interval length utilizing electronic health records. Genetics [Internet]. 2022 Dec 1 [cited 2023 Nov 2];222(4). Available from: /pmc/articles/PMC9713425/

16. Ge T, Chen CY, Ni Y, Feng YCA, Smoller JW. Polygenic prediction via Bayesian regression and continuous shrinkage priors. Nature Communications 2019 10:1 [Internet]. 2019 Apr 16 [cited 2023 Nov 2];10(1):1–10. Available from: https://www.nature.com/articles/s41467-019-09718-5

17. Seyerle AA, Sitlani CM, Noordam R, Gogarten SM, Li J, Li X, et al. Pharmacogenomics Study of Thiazide Diuretics and QT Interval in Multi-Ethnic Populations: The Cohorts for Heart and Aging Research in Genomic Epidemiology (CHARGE). Pharmacogenomics J [Internet]. 2018 Apr 1 [cited 2023 Nov 2];18(2):215. Available from: /pmc/articles/PMC5773415/

18. Al-Khatib SM, Allen LaPointe NM, Kramer JM, Califf RM. What Clinicians Should Know About the QT Interval. JAMA [Internet]. 2003 Apr 23 [cited 2023 Nov 2];289(16):2120–7. Available from: https://jamanetwork.com/journals/jama/fullarticle/1357296

19. Nachimuthu S, Assar MD, Schussler JM. Drug-induced QT interval prolongation: mechanisms and clinical management. Ther Adv Drug Saf [Internet]. 2012 [cited 2023 Nov 2];3(5):241. Available from: /pmc/articles/PMC4110870/

20. Darbar D, Kimbrough J, Jawaid A, McCray R, Ritchie MD, Roden DM. Persistent Atrial Fibrillation is Associated with Reduced Risk of Torsades de Pointes in Patients with Drug-Induced Long QT Syndrome. J Am Coll Cardiol [Internet]. 2008 Feb 2 [cited 2024 Jan 7];51(8):836. Available from: /pmc/articles/PMC2271078/

21. Johannsdottir HX, Gudmundsdottir IJ, Karason S, Sigurdsson MI. Association between pre-operative prolonged corrected QT interval and all-cause mortality after non-cardiac surgery. Acta Anaesthesiol Scand [Internet]. 2023 Mar 1 [cited 2024 Jul 22];67(3):284–92. Available from: https://onlinelibrary.wiley.com/doi/full/10.1111/aas.14178

22. Lancaster MC, Davogustto GE, Prifti E, Perret C, Funck-Brentano C, Roden DM, et al. Abstract 16962: A Polygenic Predictor of Baseline QT is Associated With Sotalol-Induced QT Prolongation. Circulation [Internet]. 2023 Nov 7 [cited 2024 Jan 8];148(Suppl_1). Available from: https://www.ahajournals.org/doi/abs/10.1161/circ.148.suppl_1.16962

23. Schwartz PJ, Woosley RL, Woosley RL. Predicting the Unpredictable: Drug-Induced QT Prolongation and Torsades de Pointes. J Am Coll Cardiol. 2016 Apr 5;67(13):1639–50.

24. Li M, Ramos LG. Drug-Induced QT Prolongation And Torsades de Pointes. Pharmacy and Therapeutics [Internet]. 2017 Jul [cited 2024 Jan 8];42(7):473. Available from: /pmc/articles/PMC5481298/

25. Li M, Ramos LG. Drug-Induced QT Prolongation And Torsades de Pointes. Pharmacy and Therapeutics [Internet]. 2017 Jul [cited 2024 Jan 8];42(7):473. Available from: /pmc/articles/PMC5481298/

26. Bihlmeyer NA, Brody JA, Smith AV, Warren HR, Lin H, Isaacs A, et al. ExomeChip-Wide Analysis of 95 626 Individuals Identifies 10 Novel Loci Associated With QT and JT Intervals. Circ Genom Precis Med [Internet]. 2018 Jan 1 [cited 2024 Jan 8];11(1):E001758. Available from: https://pubmed.ncbi.nlm.nih.gov/29874175/

27. Arking DE, Pulit SL, Crotti L, Van Der Harst P, Munroe PB, Koopmann TT, et al. Genetic association study of QT interval highlights role for calcium signaling pathways in myocardial repolarization. Nat Genet [Internet]. 2014 [cited 2024 Jan 8];46(8):826–36. Available from: https://pubmed.ncbi.nlm.nih.gov/24952745/

28. Prevarskaya N, Skryma R, Shuba Y. Ion channels in cancer: Are cancer hallmarks oncochannelopathies? Physiol Rev [Internet]. 2018 Apr 1 [cited 2024 Jan 8];98(2):559–621. Available from: https://journals.physiology.org/doi/10.1152/physrev.00044.2016

29. Bortner CD, Cidlowski JA. Ion channels and apoptosis in cancer. Philosophical Transactions of the Royal Society B: Biological Sciences [Internet]. 2014 Mar 3 [cited 2024 Jan 8];369(1638). Available from: /pmc/articles/PMC3917358/

30. Stewart TA, Yapa KTDS, Monteith GR. Altered calcium signaling in cancer cells. Biochimica et Biophysica Acta (BBA) - Biomembranes. 2015 Oct 1;1848(10):2502–11.

31. Cui C, Merritt R, Fu L, Pan Z. Targeting calcium signaling in cancer therapy. Acta Pharm Sin B [Internet]. 2017 Jan 1 [cited 2024 Jan 8];7(1):3. Available from: /pmc/articles/PMC5237760/

