## Supplementary material for "Polygenic Risk Score predicts QTc-prolongation and Short-Term Mortality in Patients using QT-prolonging Psychoactive Medications": The absolute delta-QTcF in milliseconds of ECGs measured within 18 months (table 1a) and 1 month (table 1b) before initiation of non-QT-prolonging med

**Calculation of treatment period:**

Information on the package size and the dose of the tablet/capsule were available and mean daily dose was calculated: The difference of the first and last prescription dates was defined as the treatment duration. The mass of the medication used in the treatment period was the sum of the packages in milligrams except the package of the last prescription. Division of the two latter results was used to calculate the mean daily dose.

If only one prescription had been filled, we divided the total dose of the package with the assumed daily dose to find treatment period.

**ATC-codes and assumed initial daily dose.**

QT-prolonging psychoactive medication with a known risk of Torsade de Pointes

Citalopram (N06AB04): 10 mg/day

Haloperidol (N05AD01): 5 mg/day

Escitalopram (N06AB10): 10 mg/day

Methadone (N07BC02): 5 mg/day

Chlorprothixene (N05AF03): 50 mg/day

Levomepromazine (N05AA02): 25 mg/day

Pimozide (N05AG02): 5 mg/day

Sulpiride (N05AL01): 400 mg/day

Sertindole (N05AE03): 4 mg/day

Control drugs: Non-QT prolonging psychoactive medication

Aprozolam (N05BA12): 1.5 mg/day

Bupropion (N06AX12): 150 mg/day

Chordiazepoxide (N05BA02): 10 mg/day

Duloxetine (N06AX21): 60 mg/day

Isocarboxazide (N06AF01): 10 mg/day

Vortioxetine (N06AX26): 10 mg/day

ATC-codes for medication used as covariates:

Loop-diuretics: C03C

Thiazides without potassium: C03AA

Mirtazapine: N06AX11

Nortriptyline: N06AA10

Mianserin: N06AX03

Fluanxol: N05AF01

Clozapine: N05AH02

Aripiprazole: N0AX12

Perphenazine:  N05AB03

Melperone: N05AD03

Maprotiline: N06AA21

Pipamperone: N05AD05

**ICD-codes**

Acquired long QT syndrome: ICD10:DI472F.

Drug-induced QT syndrome: ICD10: DI472FA.

Torsade de Pointes ventricular tachycardia: ICD10:DI472D.

Heart failure: ICD10: DI50, DI110, DI130, DI132, DI420, DI426, DI427, DI428 or DI429.

Ischemic Heart Disease (both ICD and procedure codes):  Myocardial infarction (ICD10: DI21, DI22, DI23, DI24, DI252, DI255, DI256, DI258, DI25, DT822 or DZ951), percutaneous coronary intervention (KFNG00, KFNG02, KFNG05, KFNG10, KFNG12, KFNG20, KFNG22, KFNG30, KFNG40, KFNG96, KZFX01) or coronary artery bypass grafting (KFNA-F, KFNH, KFNJ, KFNK, KFNW).

Hepatic insufficiency: Hepatic failure (ICD10:DK72), chronic hepatitis (ICD10:DK73) or fibrosis/cirrhosis of the liver (ICD10:DK7).

Renal insufficiency: Acute kidney failure (ICD10:DN17), chronic kidney disease (ICD10:DN18) or unspecified kidney failure (ICD10:DN19).

Cancers except non-malignant melanoma skin cancer: ICD10:DC except ICD10:DC44.

Stroke: ICD10:DI64.

COPD: ICD10:DJ44.

Atrial fibrillation or atrial flutter: ICD10:DI48

| Baseline Characteristics N=15.249 | $PRS_{QTc}$ <80 % N=12.185 | $PRS_{QTc}$ ≥ 80 %, N=3064 | P-value | | $PRS_{QTc}$ <90 % N=14.138 | $PRS_{QTc}$ ≥ 90 %, N=1537 | P-value | | $PRS_{QTc}$ <99 % N=15515 | $PRS_{QTc}$ ≥ 99 %, N=160 | P-value |
|---|---|---|---|---|---|---|---|---|---|---|---|
| **Age** | 63.28 | 63.09 | 0.25 | | 63.25 | 63.1 | 0.4 | | 63.24 | 63.4 | 0.76 |
| **Male (N, %)** | 5961 (47.9 %) | 1513 (48.27 %) | 0.73 | | 6595 (47.9 %) | 722 (48.2 %) | 0.82 | | 7230 (47.9 %) | 87 (55.76 %) | 0.06 |
| **Cancer (N, %)** | 2797 (22.9 %) | 783 (25.5 %) | 0.0026 | | 3211 (23.3 %) | 369 (24.7%) | 0.26 | | 3530 (23.4 %) | 50 (32 %) | 0.014 |
| **Stroke (N, %)** | 976 (8 %) | 276 (9 %) | 0.078 | | 1113 (8.09 %) | 139 (9.3 %) | 0.12 | | 1240 (8.2 %) | 12 (7.7 %) | 0.93 |
| **COPD (N, %)** | 1105 (9 %) | 283 (9.2 %) | 0.8 | | 1254 (9.1%) | 134 (9 %) | 0.88 | | 1367 (9 %) | 21 (13.46 %) | 0.078 |
| **Ischemic Heart Disease (N, %)** | 2334 (19.15 %) | 580 (18.9 | 0.8 | | 2629 (19.11 %) | 285 (19 %) | 0.98 | | 2887 (19.1 %) | 27 (17.3%) | 0.64 |
| **Heart Failure (N, %)** | 843 (6.9 %) | 99 (6.34 %) | 0.29 | | 939 (6.8 %) | 99 (6.6 %) | 0.8 | | 1025 (6.8 %) | 13 (8.3 %) | 0.54 |
| **Renal Insufficiency (N, %)** | 608 (5 %) | 183 (6 %) | 0.03 | | 720 (5.2 %) | 71 (4.7 %) | 0.45 | | 786 (- %) | ≤5 (- %) | - |
| **Hepatic Insufficiency (N, %)** | 137 (1.1 %) | 34 (1.1 %) | 1 | | 157 (1.1 %) | 14 (0.94 %) | 0.5 | | 169 (- %) | ≤5 (- %) | - |

| | | | | | | | | | | | |
|---|---|---|---|---|---|---|---|---|---|---|---|
| **Additional QT-prolonging drug (N, %)** | 752 (6.17 %) | 197 (6.4 %) | 0.6 | | 851 (6.2 %) | 98 (6.5 %) | 0.61 | | 9544( - %) | ≤5 (- %) | - |
| **Loop-diuretics or Thiazides (N, %)** | 836 (6.86 %) | 209 (6.8 2%) | 0.98 | | 951 (6.9 % | 94 (6.3) | 0.39 | | 1029 (6.8 %) | 16 (10.26 %) | 0.12 |

Table 1: The baseline characteristics of the sub-analysis population, in which the inclusion was not limited by before-treatment ECG and potassium blood test.

| Risk factors for mortality and prolonged QT-interval | OR for 30-day-mortality | Confidence Interval | P-value |
|---|---|---|---|
| **Age** | 1.03 | 1.03-1.04 | $<1*10^{-4}$ |
| **Male** | 1.45 | 1.21-1.74 | $<1*10^{-4}$ |
| **Cancer** | 10.61 | 8.61-13.06 | $<1*10^{-4}$ |
| **Stroke** | 1.05 | 0.78-1.42 | 0.73 |
| **COPD** | 1.61 | 1.26-2.05 | 0.00012 |
| **Ischemic Heart Disease** | 0.8 | 0.63-1.01 | 0.057 |
| **Heart Failure** | 1.13 | 0.83-1.55 | 0.43 |
| **Renal Insufficiency** | 1.68 | 1.26-2.24 | 0.0004 |
| **Hepatic Insufficiency** | 2.74 | 1.62-4.62 | 0.00015 |
| **Additional QT-prolonging drug** | 1.33 | 0.98-1.8 | 0.065 |
| **Loop-diuretics or Thiazides** | 2.13 | 1.66-2.73 | $<1*10^{-4}$ |
| $PRS_{QTc} \geq 80$ % | 1.23 | 1-1.51 | 0.053 |
| $PRS_{QTc} \geq 90$ % | 1.52 | 1.16-1.97 | 0.002 |
| $PRS_{QTc} \geq 99$ % | 1.69 | 0.87-3.26 | 0.12 |

Table 2: Results of on-treatment, 30-day-mortality in the study population of first-time users of QT-prolonging medications without baseline ECGs and potassium blood tests. In the logistic models, the risk models in the left column were included. In addition, all the models have been adjusted for PCs 1-10. N=15.675.

| 30-Day Mortality | Rest, <80 % N=12.185 | $PRS_{QTc}$ ≥ 80 % N=3064 | P-value | | Rest, <90 % N=13.764 | $PRS_{QTc}$ ≥ 90 % N=1495 | P-value | | Rest, <99 % N=15.093 | $PRS_{QTc}$ ≥ 99 % N=156 | P-value |
|---|---|---|---|---|---|---|---|---|---|---|---|
| Mortality (N, %) | 431 (3.5 %) | 137 (4.4 %) | 0.017 | | 492 (3.58) | 76 (5 %) | 0.004 | | 556 (3.58 %) | 12 (7.5 %) | 0.015 |
| Age at death (mean in y) | 73.7 | 73.9 | 0.867 | | 73.78 | 73.56 | 0.7 | | 73.67 | 75.16 | 0.567 |
| Male (N, %) | 239 (55.45 %) | 89 (64.96 %) | 0.06 | | 280 (56.9 %) | 48 (63 %) | 0.07 | | 321 (57.7 %) | 7 (58.3 %) | 1 |
| Cancer (N, %) | 341 (79.11) | 106 (77.37 %) | 0.75 | | 341 (79.11 %) | 62 (81.57 %) | 0.73 | | 435 (78.2 %) | 12 (100 %) | 0.14 |

Table 3: Results showing characteristics of the individuals deceased within 30-days after initiation of the QT-prolonging drug in the subanalysis-population without baseline ECGs and potassium blood tests.
